# Predicting Clinical Outcomes of SARS-CoV-2 Infection During the Omicron Wave Using Machine Learning

**DOI:** 10.1101/2023.08.06.23293725

**Authors:** Steven Cogill, Shriram Nallamshetty, Natalie Fullenkamp, Kent Heberer, Julie Lynch, Kyung Min Lee, Mihaela Aslan, Mei-Chiung Shih, Jennifer S Lee

## Abstract

The Omicron SARS-CoV-2 variant continues to strain healthcare systems. Developing tools that facilitate the identification of patients at highest risk of adverse outcomes is a priority. The study objectives are to develop population-scale predictive models that: 1) identify predictors of adverse outcomes with Omicron surge SARS-CoV-2 infections, and 2) predict the impact of prioritized vaccination of high-risk groups for said outcome. We prepared a retrospective longitudinal observational study of a national cohort of 192,984 patients in the U.S. Veteran Health Administration who tested positive for SARS-CoV-2 from January 15 to August 15, 2022. We utilized sociodemographic characteristics, comorbidities, vaccination status, and prior COVID-19 infections, at time of testing positive for SARS-CoV-2 to predict hospitalization, escalation of care (high-flow oxygen, mechanical ventilation, vasopressor use, dialysis, or extracorporeal membrane oxygenation), and death within 30 days. Machine learning models demonstrated that advanced age, high comorbidity burden, lower body mass index, unvaccinated status, prior SARS-CoV-2 infection, and oral anticoagulant use were the important predictors of hospitalization and escalation of care. Similar factors predicted death. However, prior SARS-CoV-2 infection was associated with lower 30-day mortality, and anticoagulant use did not predict mortality risk. The all-cause death model showed the highest discrimination (Area Under the Curve (AUC) = 0.895, 95% Confidence Interval (CI): 0.885, 0.906) followed by hospitalization (AUC = 0.829, CI: 0.825, 0.834), then escalation of care (AUC=0.805, CI: 0.795, 0.814). Assuming a vaccine efficacy range of 70.8 to 78.7%, our simulations projected that targeted prevention in the highest risk group may have reduced 30-day hospitalization, care escalation, and death in more than 2 of 5 unvaccinated patients.

## Introduction

The World Health Organization (WHO) estimates that the COVID-19 pandemic has resulted in over 521 million infections and 6.2 million deaths globally [1]. High mutation rates and the relatively rapid emergence of SARS-CoV-2 variants led to multiple surges that have strained healthcare systems worldwide. The Omicron (B.1.1.529) variant became the predominant cause of SARS-CoV-2 infections in the U.S. by January 2022 [2,3], after emerging in South Africa in November 2021 [4,5]. Although Omicron variants and sub-variants have been linked to lower rates of hospitalization and death, [3,6–8] Omicron-driven surges continued to challenge healthcare systems due to higher infectivity, partial vaccine escape, and antibody resistance [3,7].

Predictive modeling during the pandemic has provided crucial insight into clinical outcomes with COVID-19 infections; however, to date, these risk prediction tools have largely not included data for Omicron variants and have inconsistently incorporated important clinical factors such as vaccination status and prior SARS-CoV-2 infection [9–12]. In this study, we first applied machine learning (ML) models to identify baseline patient characteristics that predict risk for hospitalization, escalation of care, and mortality among SARS-CoV-2 positive US Veterans during a recent seven-month observation period (January 15 –August 15, 2022) when Omicron variants predominated. Our models incorporated previously under-utilized factors including vaccination status and prior COVID-19 infection. Then, we extended our models to quantify the predicted impact of a mitigating strategy such as prioritized vaccination of high-risk groups on reducing the short-term risk of hospitalization, escalation of care, and death during the observation period. To do this, we utilized a well-characterized cohort of U.S. Veterans with SARS-CoV-2 infection in a national Veteran Health Administration (VHA) database.

## Materials and methods

### Study cohort

Our study cohort consisted of all 192,984 Veterans who tested positive for COVID-19 between January 15 and August 15, 2022, as captured by the VHA’s COVID-19 Shared Data Resource with data curation within the VHA’s Corporate Data Warehouse (CDW). Data were accessed on 10/4/22 for research purposes. No new data were collected and no direct patient (or participant) contact took place. Patients’ curated electronic health records in the VHA’s CDW were analyzed behind the VHA secured firewall as part of the VHA research data initiative, Leveraging Electronic Health Information to Advance Precision medicine (LEAP, CSP#2012), which has been approved by VHA’s Central Institutional Review Board and Research & Development Committees at 3 VA Medical Centers (Salt Lake City, Palo Alto, and West Haven). The date of the positive test is defined as the index date. For the selected cohort within the data resource, there were no missing data for the selected fields and unknown covariates were indicated as such. Patients outside the age range of 18 to 100 or outside the Body Mass Index (BMI) range of 15 to 100 were excluded from the analysis.

### Study outcomes

We predicted the risk of developing one of the following three distinct, non-mutually exclusive clinical outcomes representing SARS-CoV-2 severity within 30 days of infection: (i) hospitalization, (ii) escalation of care (defined as the need for high-flow supplemental oxygen, mechanical ventilation, vasopressors, renal replacement therapy [with no prior dialysis in the preceding two years], or extracorporeal membrane oxygenation [ECMO]), and (iii) all-cause mortality. Patients who tested positive for SARS-CoV-2 were deemed to have ‘mild’ infection if they did not experience any of the three outcomes of interest within 30 days of infection. The Upset plot was generated using the UpsetR package [13].

### Clinical Features

A total of 159 patient characteristics including medical comorbidities, demographic data, vaccination status, prior COVID-19 infection status, and comorbidity indices were available for each patient prior to feature selection. The medical history included pre-existing conditions, procedures, and medications. All medical history values were classified using a Boolean system for presence or absence of the specific medical condition within two years prior to the current COVID-19 infection. Demographic and clinical data employed in the modeling included age, sex, race/ethnicity, blood type, BMI, veteran status, whether overweight at index date, rurality of current residence, and veteran priority status (a surrogate for income status and benefits eligibility). These covariates were multimodal (float, categorical and Boolean). Vaccination status was represented as a categorical score from 0 to 5 as follows: 0=no vaccination, 1=partial-mRNA vaccination, 2=full vaccination (two doses of mRNA or a single dose of viral vector-based vaccine) > 5 months from index date, 3=fully-vaccinated and boosted >5 months prior to the index date, 4=fully-vaccinated <5 months prior to the index date, 5=fully-vaccinated and boosted <5 months prior to the index date. Vaccines given outside of the VHA were available in the VHA COVID-19 Shared Data Resource and reflected in our dataset. Vaccination status accounted for a two-week efficacy window. A veteran was considered to be positive for prior COVID-19 infection if the prior positive test was more than two weeks before the index date of the current infection. Medical comorbidity burden was assessed by Charlson Comorbidity Index (CCI) [14] and Elixhauser Index [15] scores for the two years prior to infection. An overall CCI and Elixhauser index score was also determined. A complete list of covariates is included in S1 Table.

### Model Development and Performance

For each of the 3 main outcomes of interest, we developed a distinct binary model that incorporated 159 unique covariate features using gradient boosting automated machine learning methods. A recursive feature elimination approach was used to find the most parsimonious models. Our data was split chronologically with training/validation data from January 15, 2022 to April 15, 2022 and our test data from April 16, 2022 to August 15, 2022. Covariates with variance lower than 1% within the training set were removed, and non-binary values were scaled from 0 to 1.

Model training and optimization were performed on the training and validation sets. The H2O AI package for automated machine learning was used to train each model and the validation set was used for benchmarking the optimization process [16]. An initial heuristic search through available modeling methods using this package identified gradient boosting machines as the highest performers (data not shown). All subsequent modeling was done using gradient boosting machines. Class imbalance within this study is a bias towards patients not having a severity outcome, and this was overcome by oversampling of the minority class where patients did have a severity outcome in training of the models to allow for higher predictive performance. The binary threshold for the models was calculated by finding the threshold with the max geometric mean for specificity and sensitivity on the test set. The 95% confidence intervals for the performance metrics were determined using the stat_util python package and its bootstrapping method with 100 iterations [17].

All reported performance metrics were generated on the set aside test set. Receiver operator characteristic (ROC) and precision recall curves and their respective area under curve (AUC) were calculated using the scikit-learn metrics package [18]. The precision recall curves were normalized by using sample weights.

### Model Interpretation and Applications

Feature importance values were extracted from the H2O generated models [16]. Relative importance is calculated as the decrease in mean squared error weighted by the number of samples passing through a given node for all trees. The percentage reported here is the fraction of a given feature against all other feature relative importance values.

Shapley Additive exPlanations (SHAP) values were generated on the test set using the SHAP python package and a tree-based explainer [19]. SHAP values were calculated on random sampling of 1,000 patients from the test set. Summary plots were generated by plotting the SHAP values in a bee swarm fashion.

For simulating the impact of targeted vaccinations, we selected the unvaccinated subset of our cohort from our test set. For each strategy scenario, we projected the potential reduction in outcomes if the patients were fully vaccinated (4 score in our vaccination status). The projection required two steps. The first was to project how many symptomatic infections would be prevented and thus prevent the outcome. To accomplish this, we randomly sampled and removed patients from our target group based on a published vaccine efficacy 95% CI range of 0.708 to 0.787 which we sampled from in a uniform fashion [20]. The second was to project for the remaining patients in our target group whether being fully vaccinated would have prevented the outcome. For this we used our model and determined if their predicted outcome changed when we altered the vaccination status score from 0 to 4. We then summed the remaining outcomes in our target group to determine the reduction. The 95% confidence intervals for the projections were determined using the stat_util python package and its bootstrapping method with 100 iterations [17].

## Results

### Patient population and clinical predictors of COVID-19 infection severity

In a national VHA cohort of 192,984 Veterans who tested positive for SARS-CoV-2 during a period in which the Omicron variant predominated (January 15-August 15, 2022), the median age was 62 years and 83.8% were men (Table 1). The racial/ethnic composition of the cohort was typical for a US Veteran population; 65.2% of the patients were white, 19.5% were black, and 9% were Hispanic. Asian, Native Hawaiian or Pacific Islander, and American Indian or Alaskan Native Veterans each represented approximately 1 % of the cohort. (Table 1).

**Table 1.**
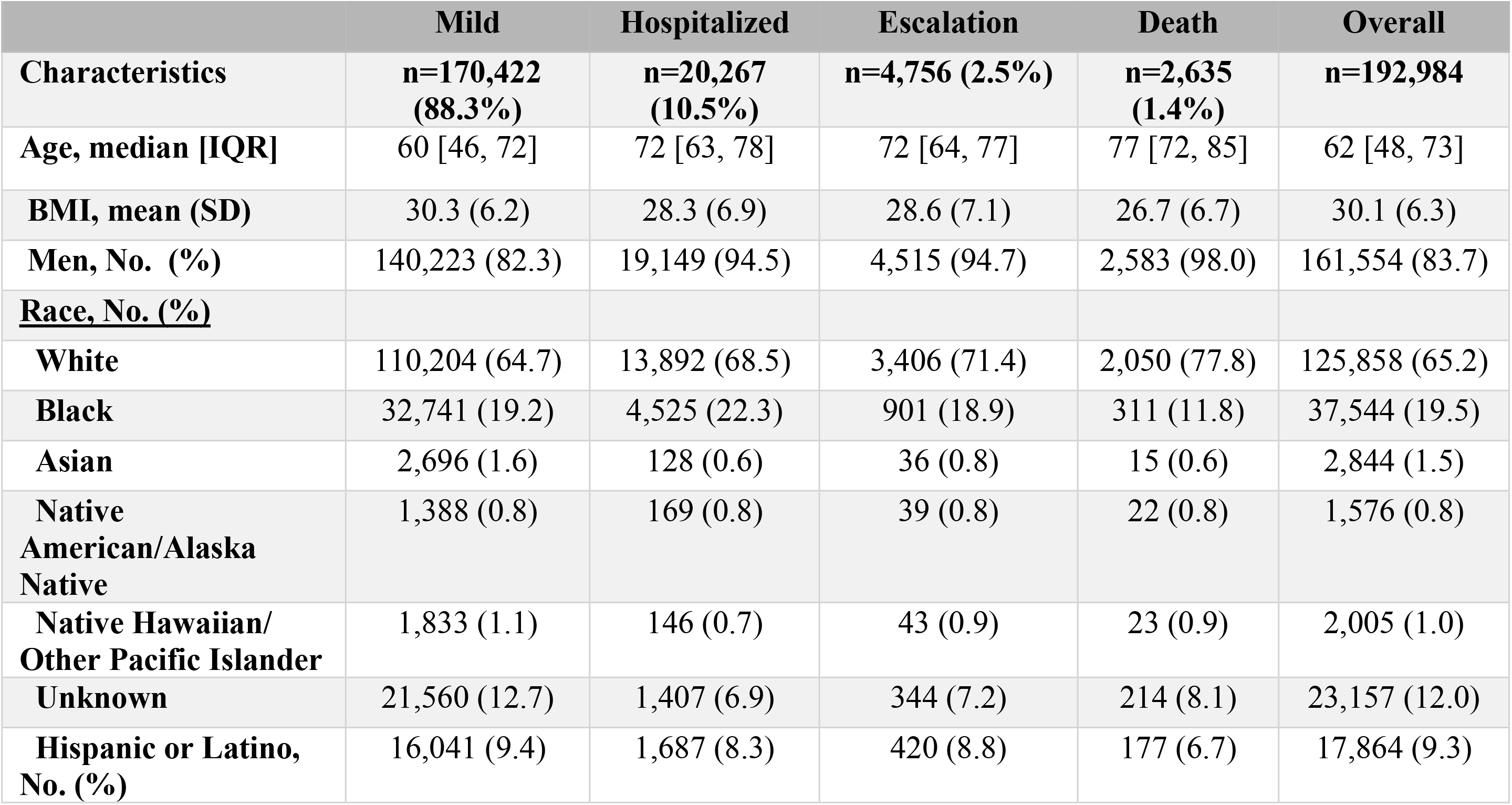
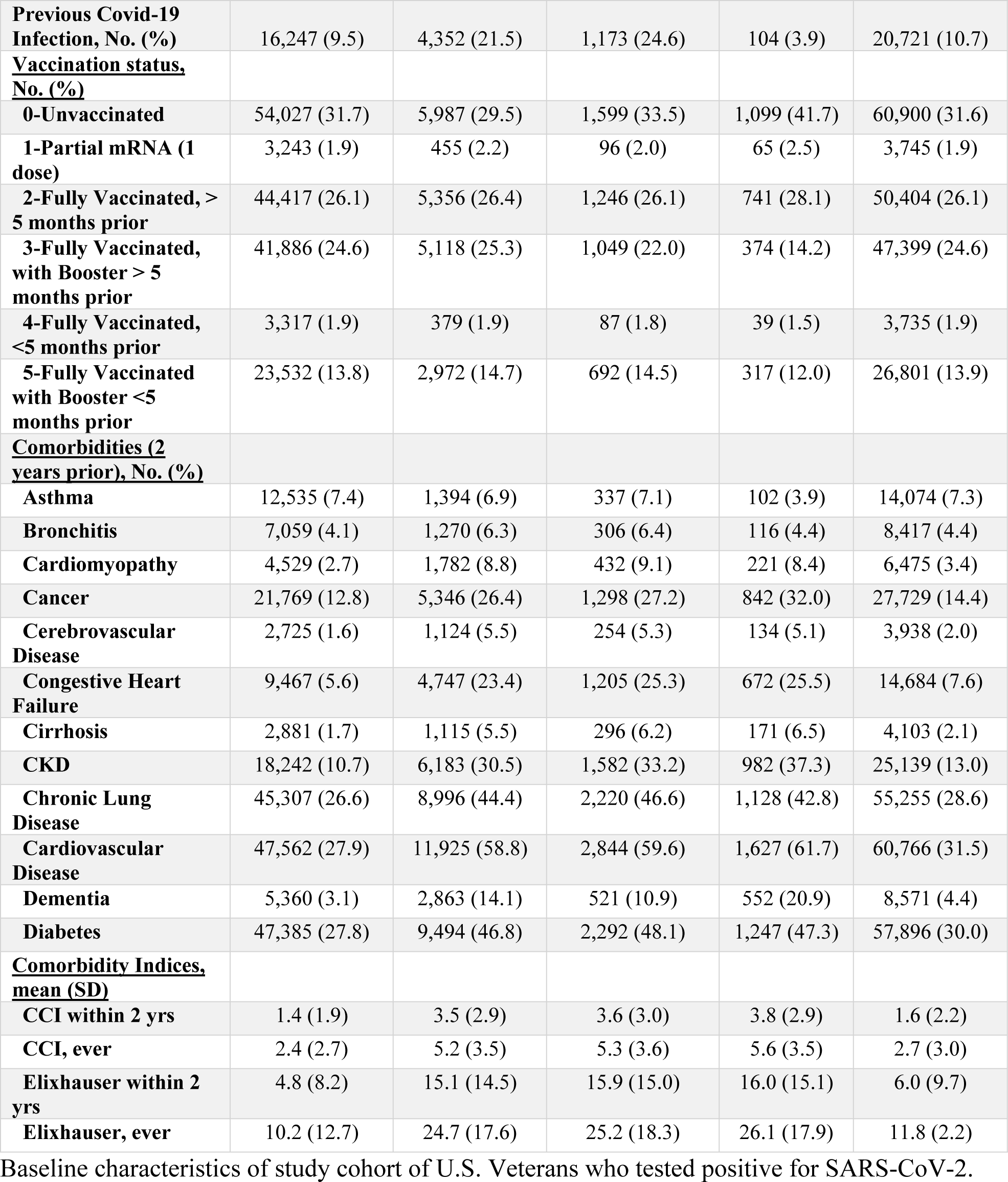
30-day outcomes after a positive SARS-CoV-2 test.

|  | Mild | Hospitalized | Escalation | Death | Overall |
| --- | --- | --- | --- | --- | --- |
| **Characteristics** | **n=170,422**<br>**(88.3%)** | **n=20,267**<br>**(10.5%)** | **n=4,756 (2.5%)** | **n=2,635**<br>**(1.4%)** | **n=192,984** |
| **Age, median [IQR]** | 60 [46, 72] | 72 [63, 78] | 72 [64, 77] | 77 [72, 85] | 62 [48, 73] |
| **BMI, mean (SD)** | 30.3 (6.2) | 28.3 (6.9) | 28.6 (7.1) | 26.7 (6.7) | 30.1 (6.3) |
| **Men, No. (%)** | 140,223 (82.3) | 19,149 (94.5) | 4,515 (94.7) | 2,583 (98.0) | 161,554 (83.7) |
| **<u>Race, No. (%)</u>** |  |  |  |  |  |
| **White** | 110,204 (64.7) | 13,892 (68.5) | 3,406 (71.4) | 2,050 (77.8) | 125,858 (65.2) |
| **Black** | 32,741 (19.2) | 4,525 (22.3) | 901 (18.9) | 311 (11.8) | 37,544 (19.5) |
| **Asian** | 2,696 (1.6) | 128 (0.6) | 36 (0.8) | 15 (0.6) | 2,844 (1.5) |
| **Native American/Alaska Native** | 1,388 (0.8) | 169 (0.8) | 39 (0.8) | 22 (0.8) | 1,576 (0.8) |
| **Native Hawaiian/ Other Pacific Islander** | 1,833 (1.1) | 146 (0.7) | 43 (0.9) | 23 (0.9) | 2,005 (1.0) |
| **Unknown** | 21,560 (12.7) | 1,407 (6.9) | 344 (7.2) | 214 (8.1) | 23,157 (12.0) |
| **Hispanic or Latino, No. (%)** | 16,041 (9.4) | 1,687 (8.3) | 420 (8.8) | 177 (6.7) | 17,864 (9.3) |
| **Previous Covid-19 Infection, No. (%)** | 16,247 (9.5) | 4,352 (21.5) | 1,173 (24.6) | 104 (3.9) | 20,721 (10.7) |
| **<u>Vaccination status, No. (%)</u>** |  |  |  |  |  |
| **0-Unvaccinated** | 54,027 (31.7) | 5,987 (29.5) | 1,599 (33.5) | 1,099 (41.7) | 60,900 (31.6) |
| **1-Partial mRNA (1 dose)** | 3,243 (1.9) | 455 (2.2) | 96 (2.0) | 65 (2.5) | 3,745 (1.9) |
| **2-Fully Vaccinated, > 5 months prior** | 44,417 (26.1) | 5,356 (26.4) | 1,246 (26.1) | 741 (28.1) | 50,404 (26.1) |
| **3-Fully Vaccinated, with Booster > 5 months prior** | 41,886 (24.6) | 5,118 (25.3) | 1,049 (22.0) | 374 (14.2) | 47,399 (24.6) |
| **4-Fully Vaccinated, <5 months prior** | 3,317 (1.9) | 379 (1.9) | 87 (1.8) | 39 (1.5) | 3,735 (1.9) |
| **5-Fully Vaccinated with Booster <5 months prior** | 23,532 (13.8) | 2,972 (14.7) | 692 (14.5) | 317 (12.0) | 26,801 (13.9) |
| **<u>Comorbidities (2 years prior), No. (%)</u>** |  |  |  |  |  |
| **Asthma** | 12,535 (7.4) | 1,394 (6.9) | 337 (7.1) | 102 (3.9) | 14,074 (7.3) |
| **Bronchitis** | 7,059 (4.1) | 1,270 (6.3) | 306 (6.4) | 116 (4.4) | 8,417 (4.4) |
| **Cardiomyopathy** | 4,529 (2.7) | 1,782 (8.8) | 432 (9.1) | 221 (8.4) | 6,475 (3.4) |
| **Cancer** | 21,769 (12.8) | 5,346 (26.4) | 1,298 (27.2) | 842 (32.0) | 27,729 (14.4) |
| **Cerebrovascular Disease** | 2,725 (1.6) | 1,124 (5.5) | 254 (5.3) | 134 (5.1) | 3,938 (2.0) |
| **Congestive Heart Failure** | 9,467 (5.6) | 4,747 (23.4) | 1,205 (25.3) | 672 (25.5) | 14,684 (7.6) |
| **Cirrhosis** | 2,881 (1.7) | 1,115 (5.5) | 296 (6.2) | 171 (6.5) | 4,103 (2.1) |
| **CKD** | 18,242 (10.7) | 6,183 (30.5) | 1,582 (33.2) | 982 (37.3) | 25,139 (13.0) |
| **Chronic Lung Disease** | 45,307 (26.6) | 8,996 (44.4) | 2,220 (46.6) | 1,128 (42.8) | 55,255 (28.6) |
| **Cardiovascular Disease** | 47,562 (27.9) | 11,925 (58.8) | 2,844 (59.6) | 1,627 (61.7) | 60,766 (31.5) |
| **Dementia** | 5,360 (3.1) | 2,863 (14.1) | 521 (10.9) | 552 (20.9) | 8,571 (4.4) |
| **Diabetes** | 47,385 (27.8) | 9,494 (46.8) | 2,292 (48.1) | 1,247 (47.3) | 57,896 (30.0) |
| **<u>Comorbidity Indices, mean (SD)</u>** |  |  |  |  |  |
| **CCI within 2 yrs** | 1.4 (1.9) | 3.5 (2.9) | 3.6 (3.0) | 3.8 (2.9) | 1.6 (2.2) |
| **CCI, ever** | 2.4 (2.7) | 5.2 (3.5) | 5.3 (3.6) | 5.6 (3.5) | 2.7 (3.0) |
| **Elixhauser within 2 yrs** | 4.8 (8.2) | 15.1 (14.5) | 15.9 (15.0) | 16.0 (15.1) | 6.0 (9.7) |
| **Elixhauser, ever** | 10.2 (12.7) | 24.7 (17.6) | 25.2 (18.3) | 26.1 (17.9) | 11.8 (2.2) |

Baseline characteristics of study cohort of U.S. Veterans who tested positive for SARS-CoV-2.

Overall, 88.3% of Veterans had mild SARS-CoV-2 infection. Among Veterans who tested positive for SARS-CoV-2, 10.5% required hospitalization, 2.5% needed escalation of care, and 1.4% died (Table 1 and Fig 1). In the subset of hospitalized infected patients, a higher percentage required escalation of care (15.3%) and died (6.0%) compared to the overall cohort (Fig 1). Patients who died or required hospitalization and/or escalation of care were older and more likely to be male. Conversely, patients who had mild infections had a higher body mass index (BMI) than those who did not (Table 1). A higher percentage of patients who died were white, compared to the overall cohort (77.8% vs 65.2%). In contrast, a lower percentage of patients who died were black, compared to those in the overall cohort (11.8% vs. 19.5%) (Table 1).

**Fig 1.**
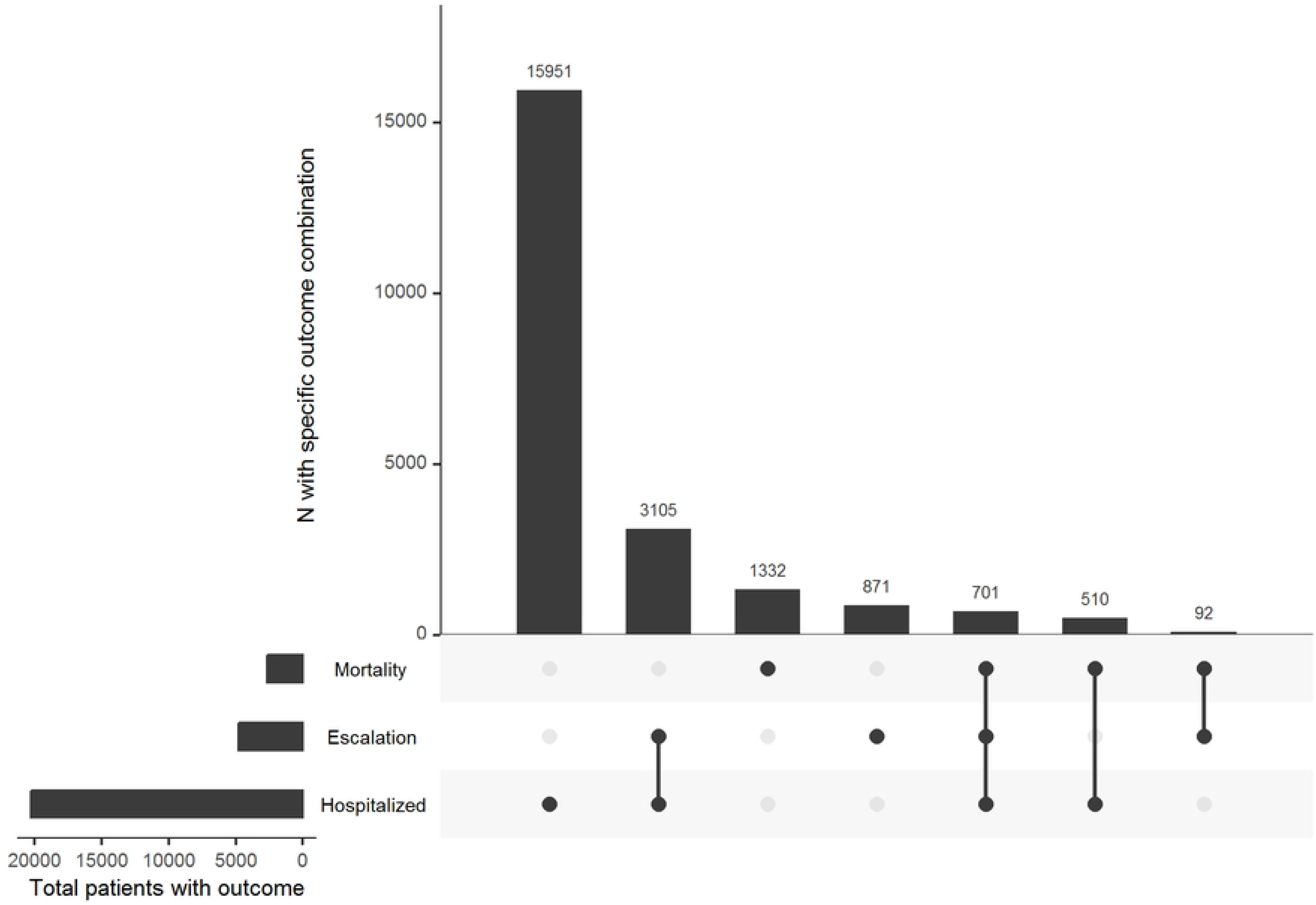
Upset plot of non-exclusive 30-day outcomes of interest in US Veterans. A dot in each row represents patients experiencing that outcome at any time within 30 days after testing positive. The vertical line connecting two (or more) dots represents patients who experienced two or more of the outcomes at any time within 30 days after testing positive.

Patients with non-mild infections had significantly higher prevalence of diabetes, congestive heart failure, cerebrovascular disease, chronic kidney disease, and cirrhosis. Dementia was also more prevalent among patients who required hospitalization, required escalation of care, or died within 30 days after testing positive. While chronic lung disease also was more prevalent, diagnoses of asthma and bronchitis in the 2 years prior to infection was not significantly different among those with any of the three outcomes of interest. The database used for this study also included information on prior SARS-CoV-2 infection as well as vaccination status (Table 1). Approximately 10.7% of the overall cohort had a history of SARS-CoV-2 infection. A higher percentage of patients with prior SARS-CoV-2 infection required hospitalization (21.5% vs 10.5% for overall cohort) or escalation of care (24.6% vs 2.5% for overall cohort). In contrast, a lower percentage (3.9%) of patients with a history of SARS-CoV-2 died within 30 days after the current SAR-CoV-2 infection.

Our study also included detailed vaccination data (Table 1). Over 31.6% of the overall cohort were unvaccinated (neither partially or fully vaccinated). Moreover, unvaccinated Veterans accounted for a disproportionately greater percentage of deaths (41.7%) compared to fully vaccinated and recently boosted (< 5 months) Veterans, who accounted for only 13% of the overall cohort and 12% of deaths. The more advanced the patients’ vaccination status, the lower their contribution to deaths (Table 1). Similar trends were observed by vaccination status for the patient groups who required hospitalization or escalation of care (Table 1).

### Model performance

After recursive feature selection evaluated the importance of 159 covariates, hospitalization had 20 relevant covariates, escalation of care had 25 relevant covariates, and mortality had 15 relevant covariates. The binary ML models predicted all 3 outcomes with good discrimination; all models had thresholds that maximized balance in performance, with sensitivity, specificity, and precision greater than 73% (Table 2). Consistent with its deterministic nature, death was predicted with better discrimination than the other outcomes, based on AUCs for both the receiver operator characteristic (ROC) (AUC = 0.895 95% CI [0.885, 0.906]) and normalized precision recall curves (AUC = 0.876 95% CI [0.867, 0.886]) (Fig 2). The model predicting hospitalization had better discrimination than the model for the need for escalation of care (hospitalization: AUC = 0.829 95% CI [0.825, 0.834]; escalated hospital care: AUC = 0.805 95% CI [0.795, 0.814]) (Fig 2).

**Table 2:** Performance of machine learning models for predicting hospitalization, escalation of care, and death within 30 days after SARS-CoV-2 infection.

| Outcome | Specificity [95% CI] | Sensitivity [95% CI] | Precision [95% CI] |
| --- | --- | --- | --- |
| **Hospitalization** | 0.73 [0.73,0.74] | 0.77 [0.76,0.80] | 0.74 [0.74,0.75] |
| **Escalation of care** | 0.74 [0.73,0.74] | 0.74 [0.72,0.76] | 0.74 [0.73,0.74] |
| **Mortality** | 0.78 [0.77,0.78] | 0.87 [0.84,0.89] | 0.79 [0.79,0.80] |

**Fig 2.**
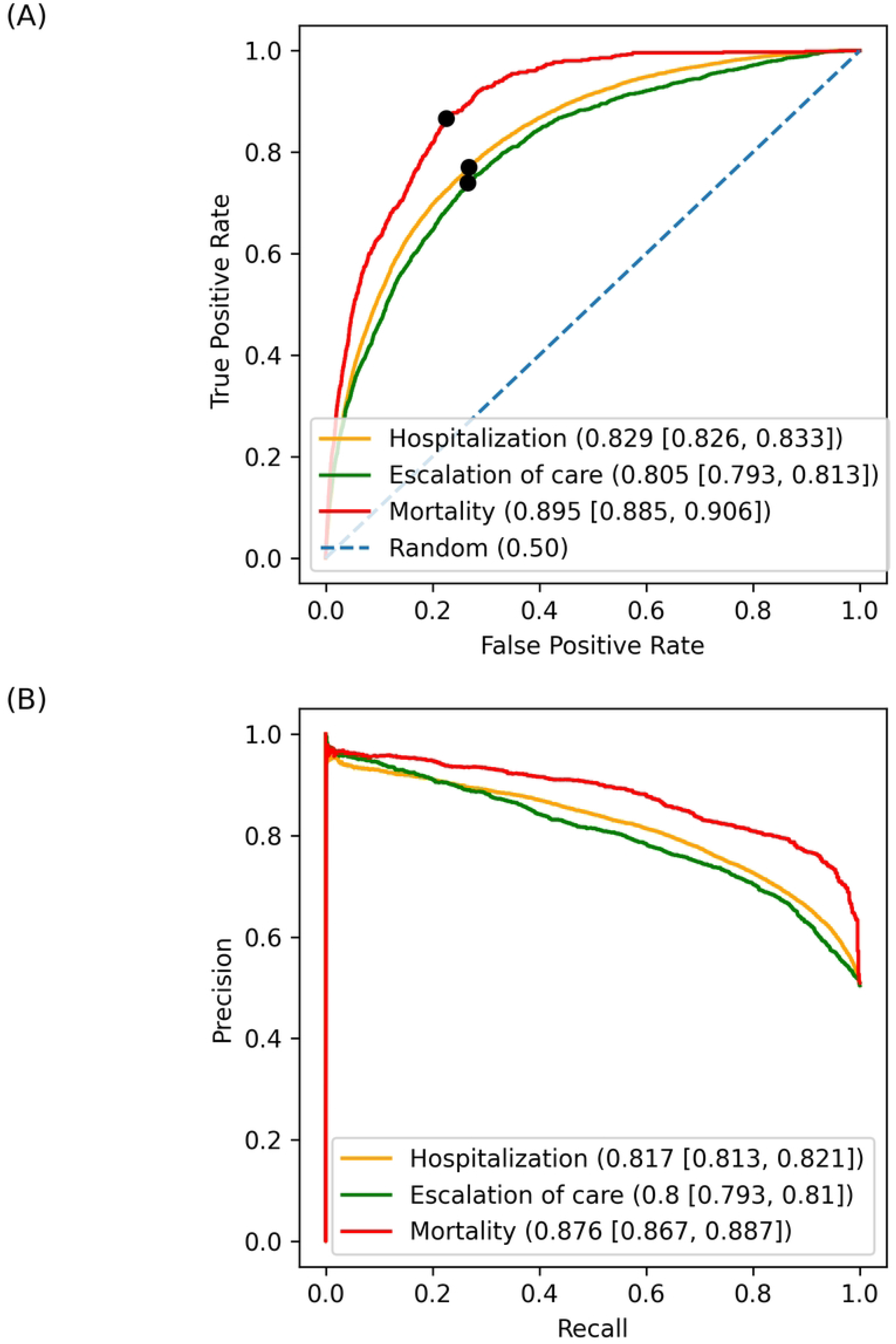
Classification performance curves with respective area under curve (AUC) and 95% confidence intervals. (A) Receiver Operating Characteristic (ROC) curve for each model with respective false positive and true positive rates at the classification thresholds indicated by black dots. (B) Normalized precision recall curve for each 30-day outcome.

### Model interpretation

We evaluated the covariates that most predicted risks of hospitalization, escalation of care, and mortality within 30 days of a SARS-CoV-2 positive test during the observation period. Feature importance was measured as the fraction of total error reduction for a given covariate (Fig 3). We generated SHAP summary plots to show the impact of covariate values on predictive output (S1 Fig). Advanced age was the second most predictive covariate for hospitalization (Fig 3A and S1 Fig A). It was also the most predictive covariate for escalation of care (Fig 3B and S1 Fig B) and mortality, accounting for more than 50% of relative importance (Fig 3C and S1 Fig C).

**Fig 3.**
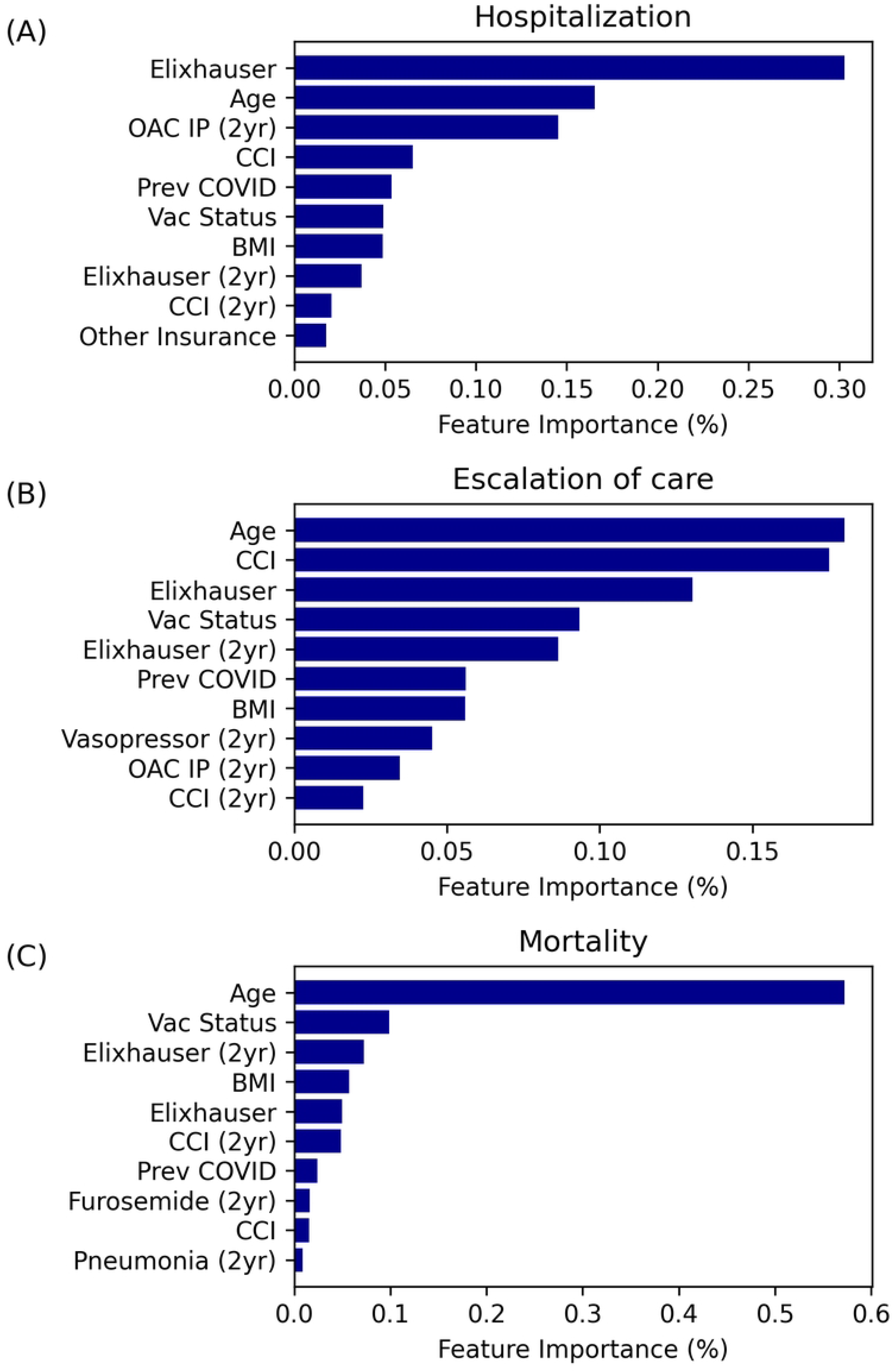
Clinical feature importance plot. (A) hospitalization, (B) escalation of care, and (C) mortality. Feature importance values for each of the three outcomes of interest are presented as a percentage, which is indicative of the fraction of error reduction that a given feature contributed to the model.

Weighted indices of comorbid illnesses, the Charlson Comorbidity index (CCI) [14] and Elixhauser index [15], were more robust predictors of the adverse outcomes than individual cardiometabolic, renal, and respiratory conditions (Fig 3). BMI was highly predictive of the outcomes; BMI was inversely proportional to predicted risk, based upon SHAP analysis (Fig 3 and S1 Fig). Veterans taking an oral anticoagulant at any time in the two years prior to testing positive for SARS-CoV-2 had higher risks of hospitalization and need for escalation of care (Fig 3A,B and S1 Fig A,B). Patients who had been prescribed vasopressors at any time in the prior two years had a higher predicted risk for escalation of care, while patients on the diuretic, furosemide, had higher predicted risk for mortality (Fig 3B,C and S1 Fig B,C).

Fully vaccinated and boosted patients had lower predicted risks of hospitalization, escalation of care, and death at 30 days. Prior SARS-CoV-2 infection predicted a lower risk of mortality but a higher risk of needing hospitalization or escalation of care (Fig 3 and S1 Fig). Additionally, unknown blood type and alternative insurance were among the most significant predictors of a lower risk for hospitalization, while a prior diagnosis of pneumonia and no acute kidney injury within two years were among the most important predictors of mortality risk (Fig 3A,C and S1 Fig A,C).

### Projected impact of risk-prioritized vaccination strategies

To project the impact of targeted vaccination on adverse outcomes using the prediction models, we examined the unvaccinated subset (n=27,903) from the test cohort (n=102,859). We projected the number of adverse outcomes for three *in silica* scenarios: (1) vaccination of all Veterans within the unvaccinated subset, (2) random vaccination of 20% of the unvaccinated Veterans, and (3) vaccination of only the Veterans in the top quintile of predicted risk for adverse outcomes (Table 3). Using sensitivity tradeoff curves (data not shown), we observed a step-up of predicted risk at the top quintile. Therefore, we selected the cut-off to be the top quintile of the population. In turn, our modeling projected the optimum impact of risk-prioritized vaccination strategy. Full vaccination of the entire unvaccinated population in our test set was predicted to reduce hospitalizations by 79% (from 2,343 to 486), escalations of care by 81% (from 470 to 87), and deaths by 82% (from 167 to 30). When a random 20% of the unvaccinated population was vaccinated in the projection modeling, hospitalizations were reduced from 2343 to 2056 (12% reduction), escalations of care from 470 to 418 (11%), and deaths from 167 to 148 (11%). When vaccinating the patients in the top quintile (20%) of the highest risk for adverse outcomes, hospitalizations were reduced from 2343 to 1353 (42%), escalations of care from 470 to 309 (34%), and deaths from 167 to 91 (45%).

**Table 3:**
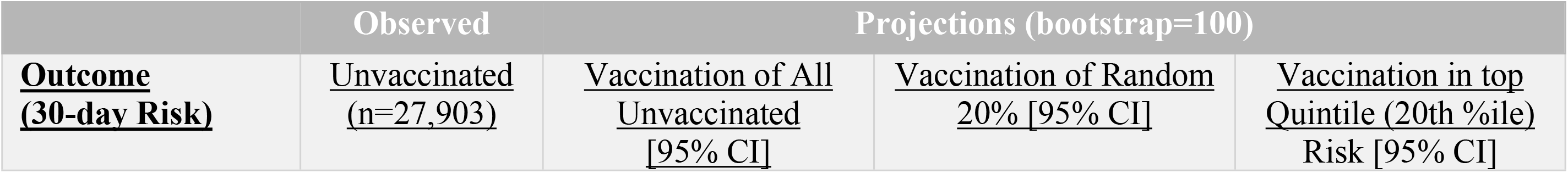
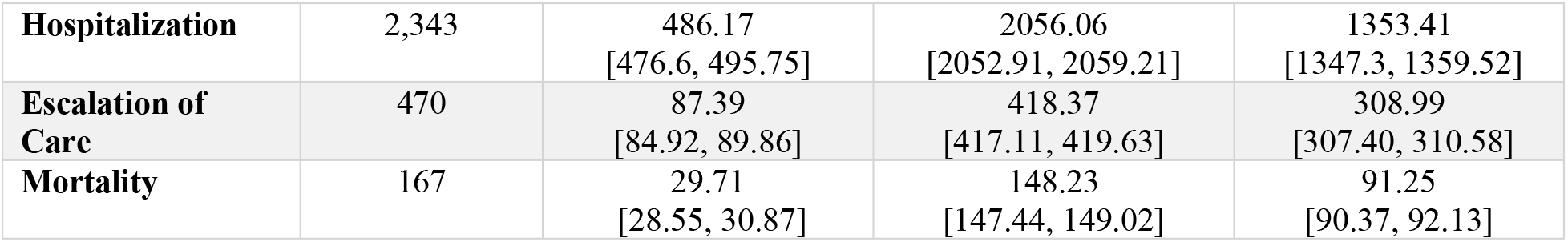
Observations and projections for occurrences for hospitalization, escalation of care, and mortality, for three vaccination scenarios.

## Discussion

In a national cohort of 192,984 US Veterans who tested positive for SARS-CoV-2 during the Omicron surge, we demonstrated the most robust prediction discrimination to date for 30-day risk for hospitalization, escalation of care, and mortality after COVID-19 infection, using ML methods. Our ML models leveraged data including detailed vaccination status and prior COVID-19 infections during the Omicron surge. We identified predictors for, and projected subgroups of, high-risk individuals who stand to benefit the most from advancing vaccination status. Prioritizing vaccination of individuals in the highest quintile of predicted risk for hospitalization or death was projected to produce greater than 3.5-fold projected reductions in hospitalization and death, compared to randomly vaccinating 20% of the population.

Previous prediction models, including those developed in the VHA, utilized data collected prior to the emergence of the Omicron SARS-CoV-2 variant [9–12]. A large retrospective analysis of over 1.5 million vaccinated patients in the VHA showed relatively low rates of breakthrough infections and related complications such as pneumonia and death [21]. This statistically powerful investigation excluded unvaccinated individuals and anyone with a prior history of COVID-19 infection, and risk prediction modeling was not a primary focus of that report. Although a prior smaller study incorporated vaccination into ML risk prediction modeling for COVID-19 [22], our study incorporated stratified vaccination status, which reflects degree of protection through number and timing of primary and booster vaccines, as well as prior infection, in an ML-driven risk prediction model.

Compared to recent studies, ML models in the present study demonstrated more robust discrimination by AUC in predicting 30-day risk for hospitalization (AUC 0.829), escalation of care (AUC 0.805), and mortality (AUC 0.895) with COVID-19 infection. Two prior studies derived from cohorts of ∼4,500 patients each demonstrated lower AUCs (0.804 and 0.813) for predicting hospitalization [23,24]. A previous model developed from a large VHA cohort of 7,635,064 (both infected and non-infected) with an observation window from May 21 to November 2, 2020 predicted 30-day mortality with a validation AUC of 0.836 (95% CI, 82.0%-85.3%) [9]. In addition, a recent study of 1,201 patients who contracted SARS-CoV-2 in Spain in 2020 predicted 30-day mortality with an AUC of 0.872 [25]. Commonly identified covariates in prior studies, advanced age and higher medical co-morbidity indices, were associated with higher risks for the adverse outcomes of interest in our models [9–11]. Our models identified a general inverse association between BMI and predicted risk for adverse outcomes. This contrasts a prior meta-analysis that demonstrated that higher BMI (and visceral adiposity) correlates with a higher risk of hospitalization, mortality, and other adverse outcomes such as admission to ICU and need for mechanical ventilation [26].

Consistent with prior vaccine trials [27], our study indicated that vaccination reduces hospitalizations, escalation of care, and deaths. Individuals who were fully vaccinated and boosted within 5 months from testing SARS-CoV-2 positive had the greatest projected protection. Importantly, our model also incorporated prior COVID-19 infection as a covariate in the risk modeling. Although patients with prior SARS-CoV-2 infection had a lower predicted 30-day mortality, they also had higher predicted risks of 30-day hospitalization and escalated hospital care. This observation is consistent with recent reports that reinfection may increase risk of any-cause mortality, hospitalization, and adverse pulmonary and extra-pulmonary health outcomes [28]. This enhanced risk of hospitalization and escalation of care is unclear but may be secondary to attendant medical comorbidities. Use of oral anticoagulants in the two years prior to current infection strongly predicted 30-day hospitalization and escalation of care. The biological basis of this observation may be related to the underlying medical conditions that warranted anticoagulation or to specific effects of the anticoagulants themselves. Notably, baseline furosemide use was also associated a higher risk of hospitalization, escalation of care and death, suggesting that underlying heart failure or volume-expanded states are important determinants of infection severity in Omicron infections.

## Limitations

The present findings in this national study of US Veterans may not be broadly applicable to the general population. Consistent with the US Veteran population, our study cohort was predominantly male and white with greater medical comorbidity and lower socioeconomic status than the general US population. The relevance of the models remains limited for racial/ethnic minority communities who have borne a disproportionate burden during the pandemic. However, the methodology used here can be applied and adapted to other populations or health care systems. For vaccine projections, all outcomes of interest were assumed to be the result of SARS-CoV-2 infection. While the VHA COVID-19 Shared Data Resource database captures all deaths, it does not capture hospitalizations and care received outside the VA. This may explain why having other non-VHA insurance was associated with lower rates of 30-day hospitalization given that patients with non-VHA insurance may have sought care outside the VA. The VHA COVID-19 Shared Data Resource database also does not establish whether SARS-CoV-2/COVID-19 is the reason for hospitalization, escalation of care, or death. Determining this is challenging. Our modeling also does not include laboratory or imaging data; these data have been shown to have robust predictive value post index date [29–32]. Finally, the model results were most relevant to Omicron variants and sub-variants and may not be relevant to other pathogenetic SARS-CoV-2 variants.

## Conclusions

Our ML risk prediction modeling approach provides robust discrimination in predicting hospitalization, escalated hospital care and death within 30 days of testing positive for SARS-CoV-2 infection during a recent observation period in which Omicron variants are the major cause of COVID-19. It can inform health care system vaccination and resource allocation decisions by characterizing individuals and subpopulations at low-to-high risk for 30-day hospitalization, escalated hospital care or death, and identifying those who might benefit least-to-most from preventive intervention. While this modeling was developed specifically for the Omicron variant surge, analogous modeling can be developed and implementable rapidly in real-time to guide vaccination strategies and resource allocation during future COVID-19 surges.

## Data Availability

Data cannot be shared publicly because it is restricted to VHA employees/users. Data are from the Corporate Data Warehouse that is part of Department of Veterans Affairs (VA), Office of Information & Technology.

## Acknowledgements

The authors would like to thank Hui Wang, Laurel Stell, and Wu Fan, for their contributions to this work.

## Supporting Information

**S1 Table. Covariates used in predictive modeling.** A table of all potential covariates that were investigated with a brief definition.

**S1 Fig.**
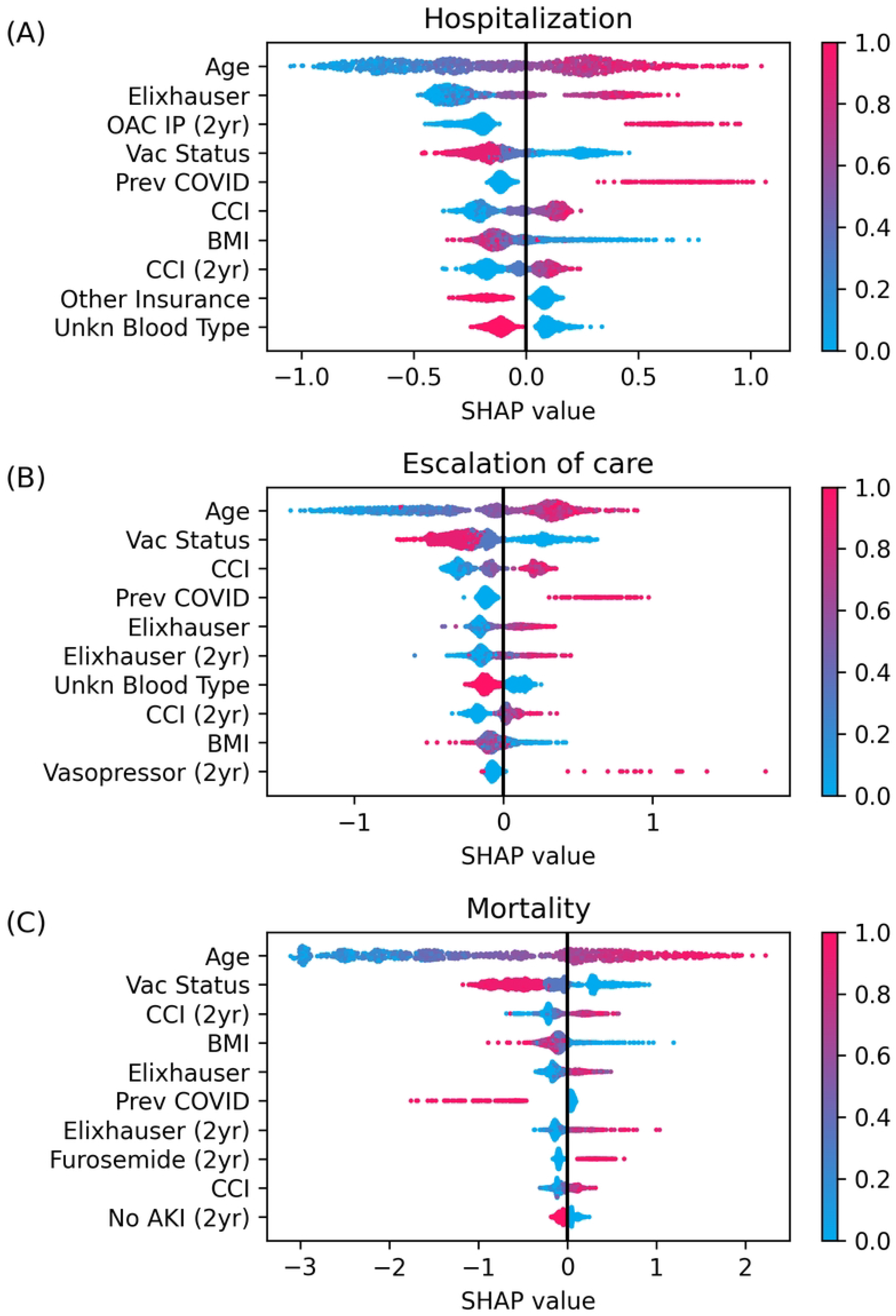
SHAP summary plots for 30-day outcomes of interest. (A) hospitalization, (B) escalation of care, and (C) mortality. Covariates are listed in order of highest to lowest impact (based on absolute mean SHAP value) along the y-axis. Each blue or red point represents a patient’s specified covariate value; that value is color coded in a heat map fashion per the legend. The x-axis is the SHAP value for the specific covariate, with SHAP values greater than 0 indicating higher predicted risk contribution and values less than 0 indicating lower predicted risk contribution for the given outcome.

## Notes

### Competing Interest Statement

The authors have declared no competing interest.

### Funding Statement

CSP#2012, Veterans Affairs All Authors

### Author Declarations

Patient clinical data were analyzed at VA Palo Alto as part of the VHA research data initiative, Leveraging Electronic Health Information to Advance Precision Medicine (LEAP, CSP#2012) research protocol, which has been approved by Institutional Review Boards and research committees at 3 VA Medical Centers (Salt Lake City, Palo Alto, and West Haven).

